# Assessment of the Publication Trends of COVID-19 Systematic Reviews and Randomized Controlled Trials

**DOI:** 10.1101/2020.08.27.20182956

**Authors:** Shunsuke Taito, Yuki Kataoka, Takashi Ariie, Shiho Oide, Yasushi Tsujimoto

## Abstract

During the COVID-19 pandemic, the number of studies listed in The National Library of Medicine registry (ClinicalTrials.gov) and preprints in medRxiv for COVID-19 has grown rapidly. In this study, we clarified the publication trends of randomized controlled trials (RCTs) and systematic reviews (SRs) regarding COVID-19. Methods: We conducted a cross-sectional study by investigating the number of SRs and RCTs on topics related to COVID-19 practice published in PubMed and medRxiv between January 1 and June 30, 2020. We calculated the ratio of the number of RCTs to that of SRs for this study period, as in a previous study. Results: The SR/RCT ratio in PubMed increased from 9.0 in March to 102 in June. In medRxiv, the SR/RCT ratio rose from 7.7 in March to 16.5 in June Discussion: The SR/RCT ratio increased and was much higher than that of 0.871 in 2017 found in a previous review of all medical research. During the study period, the trend in the COVID-19 publications comprised a more rapid increase in the number of SRs than RCTs

## Introduction

Randomized controlled trials (RCTs) and systematic reviews (SRs) are the most essential and useful study designs for evidence-based medicine and practice. The publications of SRs and meta-analyses have surprisingly increased, and the number of SRs is larger than that of RCTs in some medical fields [1]. During the COVID-19 pandemic, the number of studies listed in The National Library of Medicine’s registry (ClinicalTrials.gov) and preprints in medRxiv for COVID-19 has grown rapidly [2]. Additionally, many protocols of SRs for COVID-19 are found in PROSPERO, an international prospective register of SRs [3]. However, several reports have expressed concerns regarding unguarded, irrelevant, and misleading SRs and meta-analyses [1,4]. Therefore, in this study, we clarified the publication trends of RCTs and SRs regarding COVID-19.

## Methods

We conducted a cross-sectional study by investigating the number of SRs and RCTs on topics related to COVID-19 practice published in PubMed and medRxiv between January 1 and June 30, 2020. We excluded study protocols. The search strategies used in PubMed, which included Shokraneh’s filter for COVID-19 [5], are shown in Table 1. We retrieved abstracts from medRxiv using “random” for the RCTs, and “review,” “evidence synthesis,” “meta-analysis,” and “metaanalysis” for the SRs. Two of the four review authors (ST, YK, TA, and SO) independently selected abstracts from the filtered search results. Disagreements were resolved through discussion. If necessary, a third review author acted as an arbitrator. We calculated the ratio of the number of RCTs to that of SRs for this study period, as in a previous study [1]. A ratio greater than 1 indicated that more SRs than RCTs were published, whereas that less than 1 indicated the reverse. Ethical approval and formal consent were not required for this study.

**Table 1.**
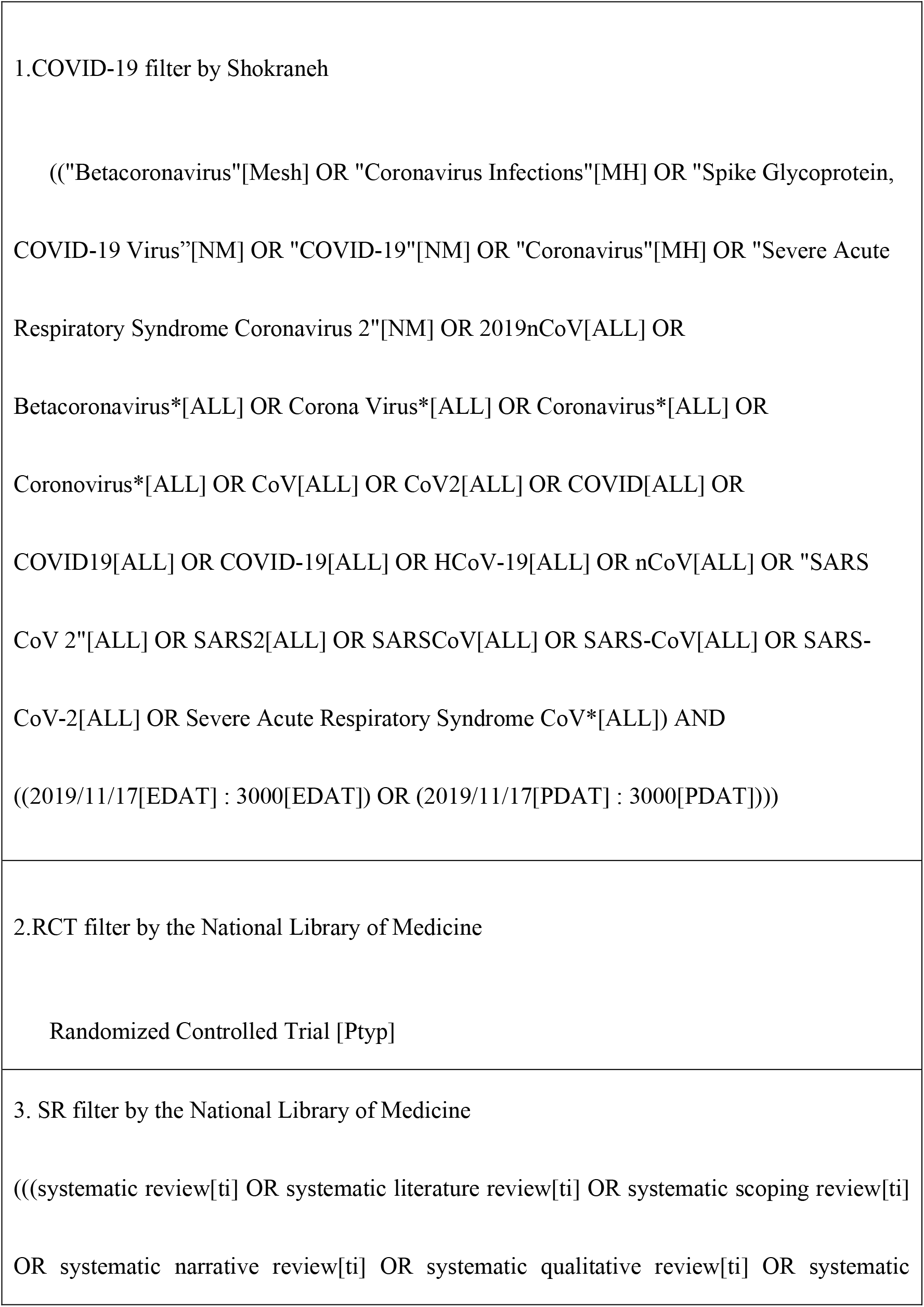
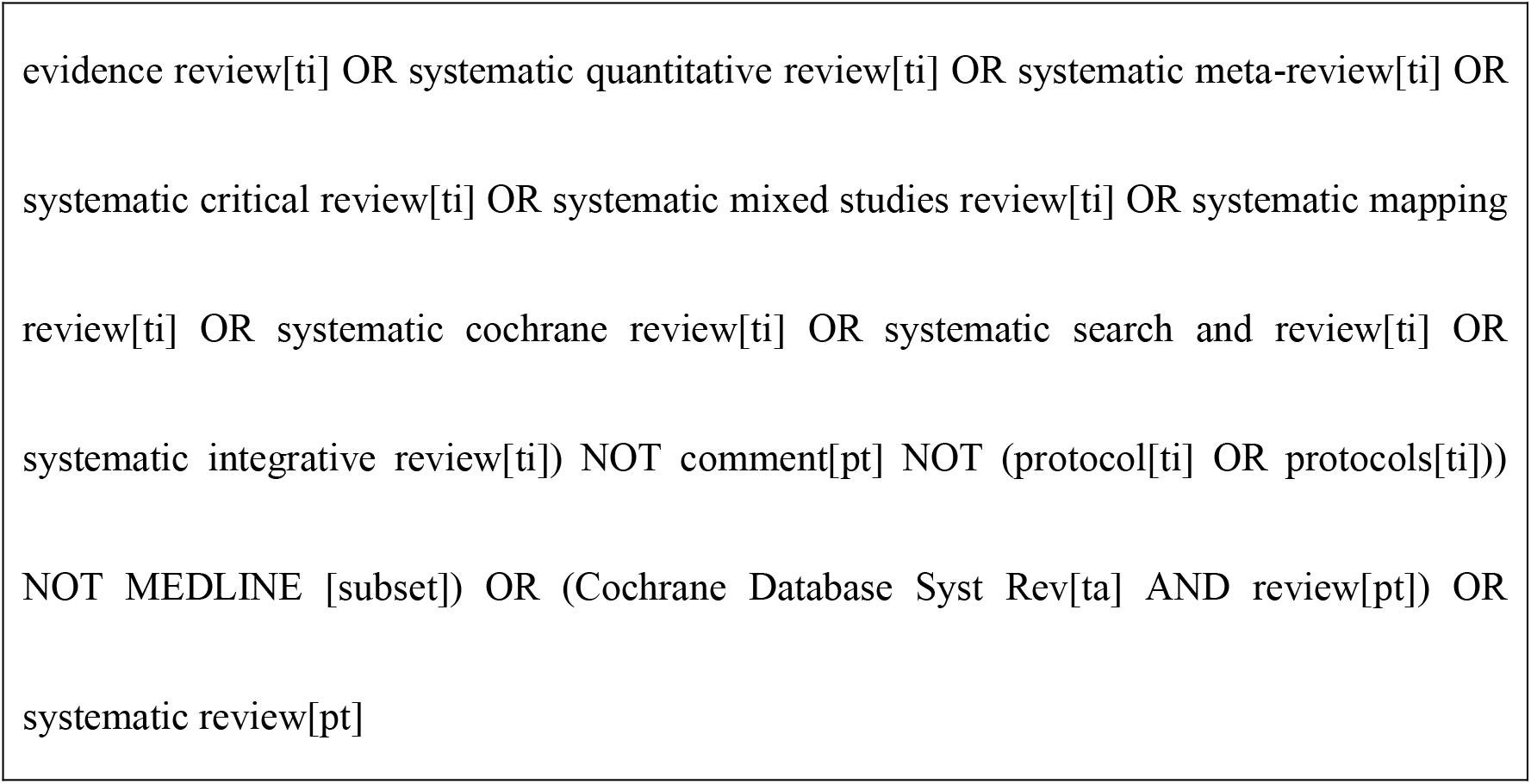

## Results

Between January and June 2020, 291 SR articles and 15 RCT articles about COVID-19 were published in PubMed, while 286 SR articles and 17 RCT articles were published in medRxiv. No RCT was published in January and February in PubMed or medRxiv. The SR/RCT ratio in PubMed increased from 9.0 in March to 102 in June. In medRxiv, the SR/RCT ratio rose from 7.7 in March to 16.5 in June (Figures 1 and 2).

**Figure 1.**
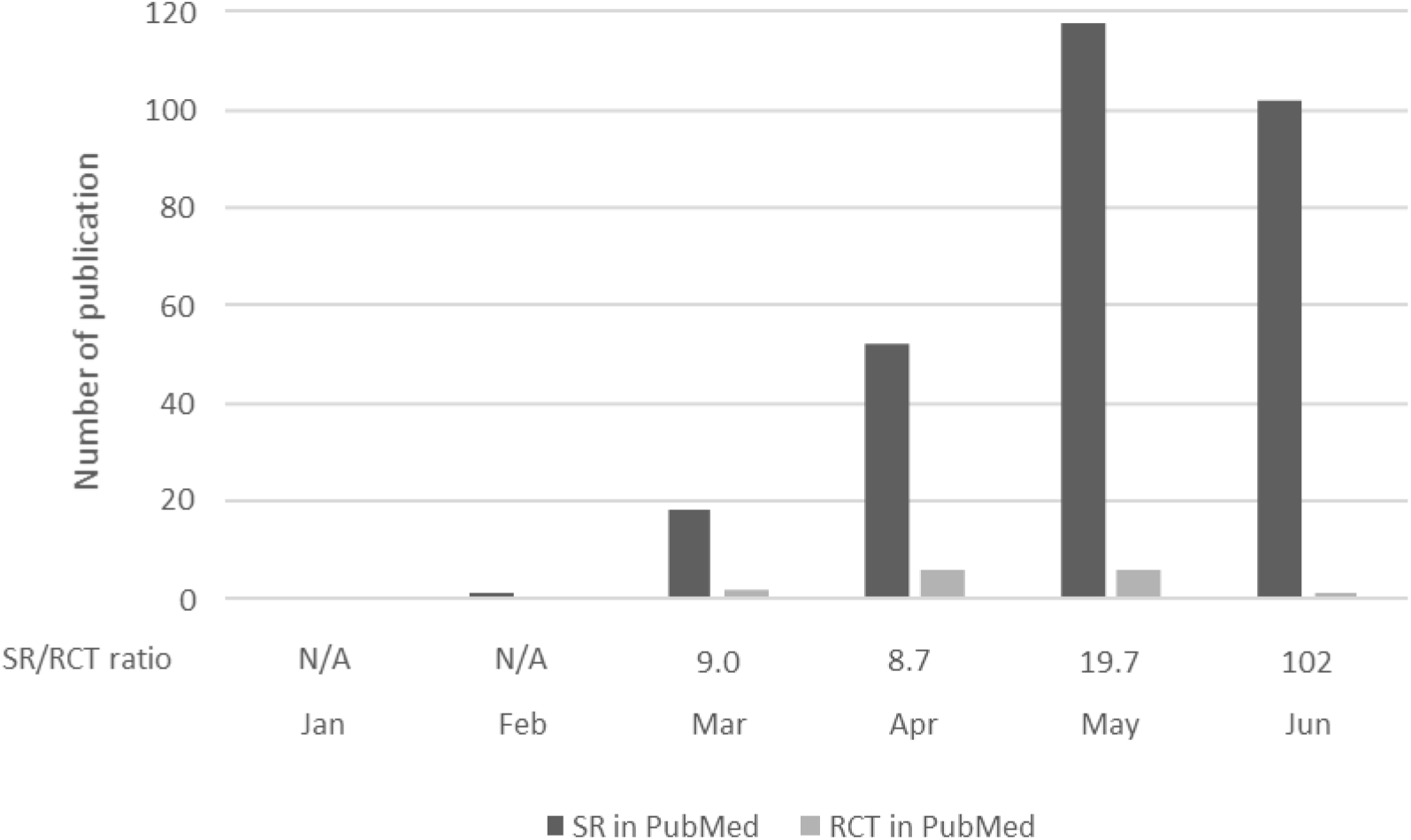
Number of PubMed systematic reviews and randomized controlled trials for COVID-19 (January to June 2020) The figure shows the trend in the number of publications of systematic reviews (including systematic reviews and meta-analyses) and randomized controlled trials for COVID-19. It also presents the ratio of systematic reviews to randomized controlled trials for COVID-19. A ratio greater than 1 indicates that more systematic reviews than randomized controlled trials were published, whereas that less than 1 indicates that more randomized controlled trials than systematic reviews were published. N/A means “not available” and indicates that the number of published systematic reviews was zero. SR = systematic review; RCT = randomized controlled trial; N/A = not available.

**Figure 2.**
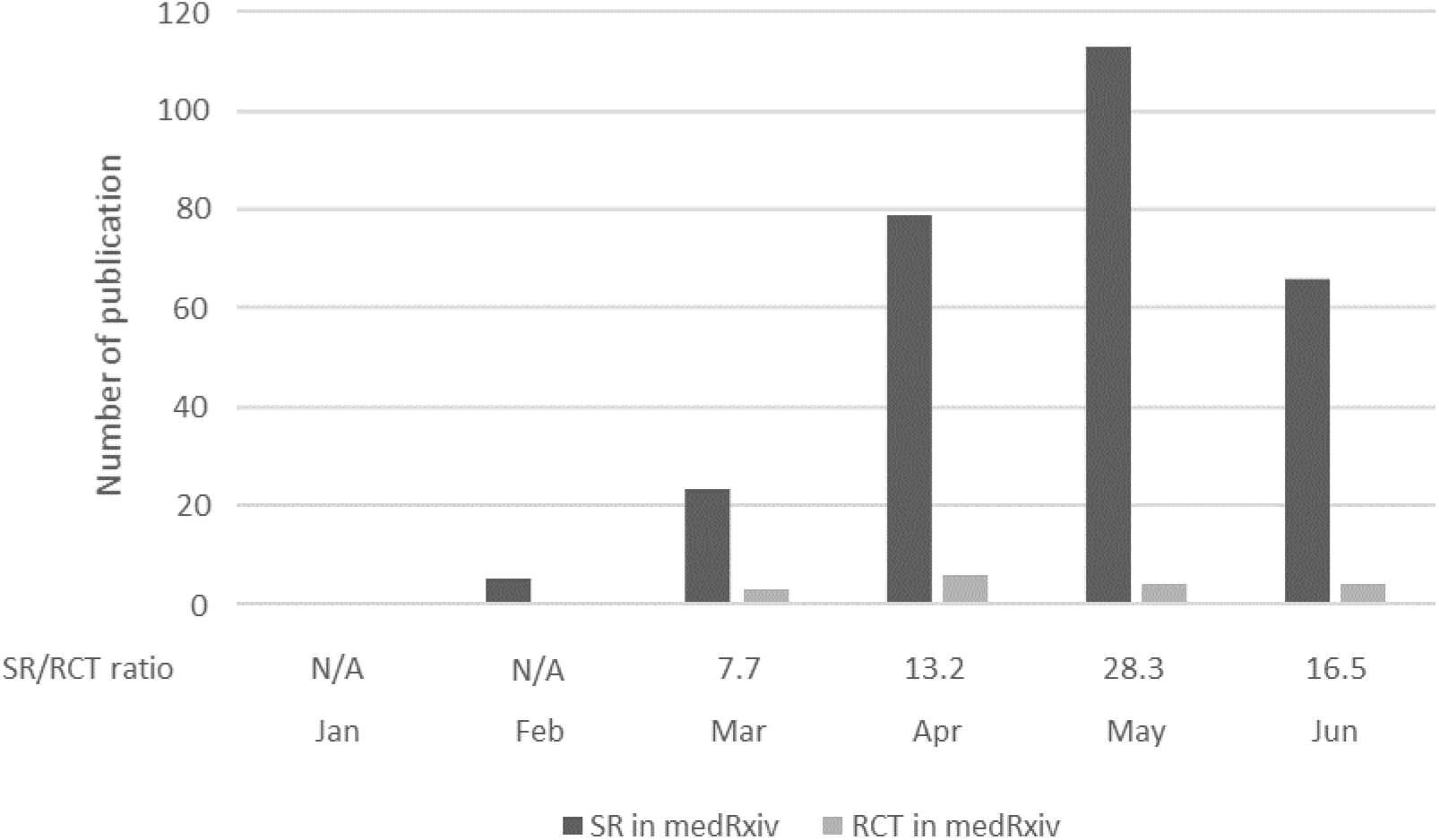
Number of medRxiv systematic reviews and randomized controlled trials for COVID-19 (January to June 2020) The figure shows the trend in the number of publications of systematic reviews (including systematic reviews and meta-analyses) and randomized controlled trials for COVID-19. It also presents the ratio of systematic reviews to randomized controlled trials for COVID-19. A ratio greater than 1 indicates that more systematic reviews than randomized controlled trials were published, whereas that less than 1 indicates that more randomized controlled trials than systematic reviews were published. N/A means “not available” and indicates that the number of published systematic reviews was zero. SR = systematic review; RCT = randomized controlled trial; N/A = not available.

## Discussion

This is the first report about the publication trends of SRs and RCTs for COVID-19. The SR/RCT ratio increased and was much higher than that of 0.871 in 2017 found in a previous review of all medical research [1]. A limitation of this study was that when we counted the number of SRs, we included non-interventional SRs, as in a previous study [1]. Although the optimal SR/RCT ratio has not been determined, this mass production of SRs could be harmful [4]. In conclusion, during the study period, the trend in the COVID-19 publications comprised a more rapid increase in the number of SRs than RCTs. Researchers may benefit from using their research resources for something other than an SR.

## Data Availability

All data associated with this manuscript are included in
the main text and supplementary materials.

## Acknowledgements

The authors would like to thank Editage (http://www.editage.jp) for English language editing.

